# Comparative LUSZ Therapeutic Study (LUSZ_AVIST) of Antiviral, Antiretroviral, and Immunosuppressive Treatments in Hospitalized COVID-19 Patients with High-Risk Factors, Biomarkers, and Disease Progression

**DOI:** 10.64898/2026.04.10.26350587

**Authors:** Elisa Makdessi, Fadi Fenianos, Nadine Nasreddine, Wissame Daher, Samar El-Hamoui, Nehman Makdissy

**Author notes:** Corresponding author: Prof. Nehman Makdissy; Lebanese University, Faculty of Sciences III, Ras-Maska, Lebanon.

## Abstract

COVID-19 has spread rapidly and caused a global pandemic making it one of the deadliest in history. Early identification of patients with coronavirus disease 2019 who may develop critical illness is of immense importance. Therefore, novel biomarkers were needed to identify patients who will suffer rapid disease progression to severe complications and death. Many treatments were adopted including the antiviral Remdesivir, the antiretroviral Lopinavir /Ritonavir and Tocilizumab. Our study aimed not only to specify high-risk factors and biomarkers of fatal outcome in hospitalized subjects with coronavirus but also to compare the efficacy of the three considered treatments to help clinicians better choose a therapeutic strategy and reduce mortality. We divided the population (n=711) into four main groups based according to the WHO ordinal severity scale. The percentage of mortality, in and out the hospital, the length of stay in the hospital, the pulmonary inflammatory lesion and its distribution, the SARS-CoV-2 IgM and IgG variations at admission, the inflammatory markers, the complete blood count, the coagulation factors and enzymes, proteins and electrolytes profile, glucose and lipid profile, and other relevant markers were measured. The significance of the observed variation was assessed by multivariate and ANOVA analyses. We succeeded to establish a novel predictive scoring model of disease progression based on a cohort of Lebanese hospitalized patients relying on the pulmonary inflammatory lesions, inflammation biomarkers such as LDH, D-Dimer, CRP, IL-6 and the lymphocyte count, the number of comorbidities and the age of the patient which all were significantly correlated with the illness severity showing best outcomes with immunomodulatory and anticoagulant treatments by the results. As top tier, Tocilizumab was more efficient than the two other treatments in non-severe cases but none of the used treatments was insanely effective alone to reduce mortality in severe cases.

## 1 Introduction

Coronavirus disease 2019 (COVID-19) has spread rapidly and caused a global pandemic, as defined by the WHO, within a short period of time. Early identification of patients with COVID-19 who may develop critical illness is of great importance. The scientific community is in urgent need of reliable biomarkers related to COVID-19 disease progression, in order to stratify high risk patients. The rapid disease spread necessitates the immediate categorization of patients into risk groups following diagnosis, to ensure optimal resource allocation. Novel biomarkers are needed to identify patients who will suffer rapid disease progression to severe complications and death. The identification of novel biomarkers is strictly related to the understanding of viral pathogenetic mechanisms, as well as cellular and organ damage. Effective biomarkers would be helpful for screening, clinical management, and prevention of serious complications. Some hematological parameters, including white blood cell (WBC), lymphopenia, c-Reactive Protein (CRP), and some biochemical parameters, such as lactate dehydrogenase (LDH), creatine kinase (CK), and troponin were reported to be associated with COVID-19 severity. [1,2]

Indeed, many previous studies suggested and developed predictive scores that estimate the severity of the disease, the risk of needing an invasive mechanical ventilation (IMV) among patients with COVID-19 and the risk of mortality; many scores including so many relevant parameters have been used to predict the status of a patient. In fact, having a high risk of death was significantly correlated to the following parameters: elder age (around 71 years old), suffering from comorbidities especially more than two, saturation in oxygen about 93% or less, NLR about 6.25 as a cut-off (high neutrophil & low lymphocyte counts), low albumin levels (about 37g/L as a cut-off), low platelets count (about 155×10^9^ /L as a cut-off), according to the CALL, Chosen, HA2T2, ANDC, ABC2-SPH, ICOP, SEIMC,IRS, NLP, APACHE II, & GRAM risk scores. Many prognostic models have been introduced to guide treatment and resource management. However, data on the impact of Comorbidity-Age-Lymphocyte count-Lactate dehydrogenase (CALL) score and Respiratory Assessment Scoring (RAS) model in predicting disease progression and mortality in COVID-19 are scarce. [3] Previous studies have demonstrated that patient’s initial laboratory analysis with high neutrophil count (>0.7 × 103/μL), lymphopenia (<0.8 × 103/μL), elevated CRP (>4.75 mg/dL), and elevated LDH (>593 U/L) were the most important predictors of mortality. [4] Another prognostic marker the lymphocyte-to-CRP ratio (LCR), was also helpful. A rise in the NLR and decline in LCR correlates with the severity of COVID-19. Specifically, a low LCR at presentation was seen to predict ICU admission and need for invasive ventilation. Patients critically ill with COVID-19 had a hyperinflammation, the associated biomarkers may be beneficial for risk stratification. Many studies found the association between several biomarkers, including serum CRP, procalcitonin (PCT), D-dimer, and serum ferritin, and COVID-19 severity. [5] Subsequent studies demonstrate that the inflammatory response plays a critical role in COVID-19, and inflammatory cytokine storm increases the severity of COVID-19. In fact, the serum levels of IL-6 can effectively assess disease severity and predict outcome in patients with COVID-19. [6]

Concerning treatments, antivirals, antiretrovirals, immunosuppressors, anti-inflammatory, corticotherapy and other were adopted; among them, the antiviral Remdesivir which is an adenosine analogue that can target the RNA-dependent RNA polymerase and block viral RNA synthesis [7], the antiretroviral Ritonavir which was recommended by the National Institutes of Health’s COVID-19 guidelines, it’s a protease inhibitor since the proteases encoded by most viruses play a crucial role in the viral life cycle. Protease inhibitors block the final step of virion assembly, this inhibition results in the production of immature virus particles [7] and the potential therapeutic IL-6 inhibitor Actemra (Tocilizumab) which is the first marketed IL-6 blocking antibody through targeting IL-6 receptors, it calms the inflammatory storm through blocking IL-6 receptors thus reducing the mortality of severe COVID-19 [8] Our distinct study aimed not only to specify high-risk factors and biomarkers of fatal outcome in hospitalized subjects with coronavirus depending on many variables analysis, but also to compare the efficacy of three different treatments (antiviral, antiretroviral, & immunosuppressive IL-6 receptor antagonist) in order to help clinicians better choose a therapeutic strategy. For it we succeeded to establish a novel predictive scoring model of disease progression based on a cohort of Lebanese patients hospitalized at Saydet zgharta hospital including those who had comorbidities such as obesity, cardiovascular diseases, and other diseases, those who needed an intensive care, and those who were oxygenated & treated with different medicines. Using this novel score model, clinicians can predict which patients admitted to the hospital with COVID-19 will need admission to an intensive care unit, mechanical ventilation, or will die; this may help identify patients with COVID-19 who may subsequently develop critical illness. Thus, we can improve the therapeutic effect and reduce the mortality of COVID-19 with more accurate and efficient use of medical resources, it helps delivering proper treatment according to each case.

## 2 Methods

### 2.1 Study Design

This study is based on data of hospitalized patients collected and followed from March 28, 2020, during a 5-year period, from the Saydet Zgharta University Medical Center (SZUMC). Patients with COVID-19 were diagnosed by polymerase chain reaction (PCR) test taken from a nasopharyngeal sample, throat sputum, saliva, urine, stool, or bodily fluid. Analyses were realized upon admission as well 8-10 days after admission. The collection of data from each patient in terms of laboratory data, treatments, and outcomes were verified by the principal investigator through the review of clinical records. Selected patients were divided into groups according to the WHO ordinal clinical severity scale (WOSS).

### 2.2 Ethics statement

All clinical investigations on human samples were conducted according to the principal expressed in the Declaration of Helsinki, as revised in 2008 (http://www.wma.net/e/policy/b3.htm). All donors provided written informed consent, and samples were collected accordingly to the ethical codes: the study protocol was approved by the institutional review committee of the SZUMC (MA-LE-E-60/2022).

### 2.3 Eligibility Criteria for Hospitalised Patients

#### As inclusion criteria

(1) admission to hospital, (2) fulfills WHO case definition, including a positive PCR for COVID-19 from any specimen (e.g., nasopharyngeal, throat, saliva, urine, stool, other bodily fluid), (3) not received any therapy (radiotherapy, chemotherapy, corticotherapy, hormonotherapy, immunotherapy, anti-inflammatory, antibiotics, antiparasitic, antiviral, antibacterial, convalescent plasma, monoclonal antibodies, or other treatments such as hydroxychloroquine and azithromycin) before admission and samples’ collection, and (4) Spo2 < 90%.

#### As exclusion criteria

(1) Non-SARS-CoV-2, (2) active indication and use for one of the investigational products (e.g., HIV positive if antiretroviral agents were used), (3) allergy or hypersensitivity to one of the investigational products (Lopinavir/Ritonavir, Remdesivir, Tocilizumab) or other contraindication, (4) progression to death is imminent and inevitable within the next 24 hours, irrespective of the provision of treatments, (5) received any therapy (radiotherapy, chemotherapy, hormonotherapy, immunotherapy, anti-inflammatory, antibiotics, antiparasitic, antiviral, antibacterial, convalescent plasma, monoclonal antibodies, or other treatments such as hydroxychloroquine and azithromycin) before admission and samples’ collection, (6) weight loss during the last 2 years, (7) abdominal surgeries, (8) pregnancy, and (9) SpO2 ≥ 90%, and (10) vaccinated individuals were excluded.

### 2.4 Treatments Prescribed: Comparative Arms

All patients received Standard Care (SC) enhanced by corticotherapy (corticosteroid, the methylprednisolone, for its anti-inflammatory and immunosuppressive effects): named here CTSC protocol. Patients were treated with or without antiviral, antiretroviral (Remdesivir, Lopinavir/Ritonavir) or immunosuppressive anti-IL-6R (Tocilizumab). (Supplementary table 1)

Three comparative groups were studied as indicated in the following table:

|  | Control | A | B |
| --- | --- | --- | --- |
| Antiviral, antiretroviral (Remdesivir, Lopinavir/Ritonavir) | (-) | (+) | (-) |
| Immunosuppressive anti-IL-6R (Tocilizumab) | (-) | (-) | (+) |
| CTSC | (+) | (+) | (+) |

### 2.5 Serology and Blood Laboratory Analysis

Blood samples were obtained from patients and analyzed in accordance with the standard tests and laboratory operating procedures. A list of assessments was done on complete blood or serology: Urea, Uric Acid, Albumin, Globulin, Total Bilirubin, Direct Bilirubin, Total Proteins, Creatinine, EGFR, Sodium, Potassium, Chloride, Calcium, Vit-B12, Vit-D, Blood sugar, HbA1c, T-Chol, HDL, LDL, TG, PT, aPTT, D-dimer, Fibrinogen, Troponin, ATIII *(anti-thrombin 3),* Protein C, Protein S, LDH, CRP, IL-6, Iron, Ferritin, CPK, C3, C4, ALT (SGPT), AST (SGOT), ALP, GGT, Lipase, Amylase, Hemoglobin, Hematocrit, RBC, WBC, Platelets, Neutrophiles, Lymphocytes, Monocytes, Eosinophils, Basophils, ESR. The NLR *(Neutrophil to Lymphocytes Ratio),* CAR (CRP to Albumin Ratio) ratios were calculated. For viral assays, whole blood samples in EDTA tubes were collected for RNA assays, and serum for IgG/IgM assays.

### 2.6 Quantitative Viral RNA Testing by RT-qPCR

Nasal swab sampling (Nasopharyngeal, Nostril, or other types of collection) for study-related viral monitoring was performed using the approved CE-IVD KITS: QIAamp Viral RNA Mini Kit (Ref. 52904, QIAGEN) for viral RNA isolation, and ANDIS® SARS-COV-2 RT-QPCR DETECTION KIT (Ref. 3103010069, 3DMED) for viral detection following guidance from the manufacturers.

### 2.7 Quantitative Viral IgG and IgM Testing: A Serology-Based Immunoassay

Accompanying an antibody test assessing immunoglobulin class G and M (IgG and IgM) with an RNA test improves overall sensitivity of the viral diagnosis in the early stage of the infection. SARS-CoV-2 IgG and IgM antibodies were assessed serologically using SARS-CoV-2 IgG and SARS-CoV-2 IgM kits following the manufacturer’s instructions (Abbott, UPO, Lebanon). The presence of IgM antibodies allows for the identification of recent infection and evaluation of disease course. The persistence of IgG antibodies allows identification of patients who have been infected in the past or recovered from the illness. 3 phases can be distinguished where the patient is positive for: (1) IgM alone (up to 7 days of infection: D0-D7, approximately), (2) IgM+IgG (∼ 30 days: D7-D35) and (3) IgG alone developing immune response (> D35).The SARS-CoV-2 IgM/IgG assays are chemiluminescent micro particle immunoassay (CMIA) used for the detection of IgM antibodies to SARS-CoV-2 in human serum and plasma on the Alinity i and ARCHITECT i Systems. These results provided by Abbott suggested that the cutoffs for IgM and IgG were 1; above 1, detection is considered positive. The manufacturer reported clinical sensitivity of 99.63% for IgG and 99.56% for IgM. An in-house validation study, using a set of sera from 25 negative controls and 25 RT-PCR-confirmed SARS-CoV-2 infection cases was conducted to estimate test performance. This validation pilot study revealed sensitivity of 95.3% and a specificity of 100%.

### 2.8 Statistical analysis

Excel 2019, SPSS and SAS match, and GraphPad Prism 10 were used for statistical evaluations. The results were expressed as mean ±SEM or percentage and analyzed for statistical significance using Student’s t-test. Logistic regression analysis with adjustment for relevant significant variables, was used to assess the frequencies and statistically analyzed using Odds ratio (OR) with 95% confidence intervals. OR>1 indicates increased occurrence of the risk factor; OR<1 indicates decreased occurrence of the risk factor (protective role). The differences in mortality were shown graphically using Kaplan-Meier curves with their log-rank test (event: death; censored data: hospital discharge). Differences between groups were determined using 1-way analysis of variance (ANOVA). For all statistical tests, α and *P* values were two-tailed and the level of significance was set at 0.05.

## 3 Results

### 3.1 Characteristics of the studied population

Our study regroups 711 unrelated subjects of white race, hospitalized at Saydet zgharta hospital, diagnosed with COVID-19 at admission, with mean age 62.4 ± 0.67 years old (60.9% males & 39.1% females) (**Table 1**)

**Table 1:** Characteristics of the studied population.

|  | *n* | % |
| --- | --- | --- |
| **COHORT** | 711 |  |
| Number of patients | 711 |  |
| Age, years (Mean ± SEM) | $62.4 \pm 0.67$ | |
| Gender (males; females) | 433; 278 | 60.9; 39.1 |
| Race, white | 711 | 100 |
| Diagnosed COVID-19 at admission (PCR positive for SARS-CoV-2) | 711 | 100 |

### 3.2 Primary outcomes and comorbidities

**Table 2** shows that 118 patients out of 711 (16.6%) had no comorbidities, 25.88% patients had only one comorbidity, 22.64 % had 2 comorbidities, while 18.57 % suffered of 3. This population includes mainly hypertensive patients (344 patients out of 711 (48.4%) had arterial hypertension). 42.3% of the patients were obese (BMI greater than 30), while 31.7% had type II diabetes. Concerning the mortality, this table shows that 147 patients out of 711 (20.7%) died. All of them died in the hospital and 88 patients out of 711 (12.4%) died in the intensive care unit ICU.

**Table 2:** Primary outcomes and comorbidities.

|  | *n* | % |
| --- | --- | --- |
| **COMORBIDITIES** (*listed in descending order of occurrence*) |  |  |
| Number of comorbidities, *n* (Mean ± SEM, [range]) | $2 \pm 0.05$, [0-7] | |
| Number of comorbidities per patient, category (*n*, %) |  |  |
| 0 | 118 | 16.60 |
| 1 | 184 | 25.88 |
| 2 | 161 | 22.64 |
| 3 | 132 | 18.57 |
| 4 | 77 | 10.83 |
| 5 | 32 | 4.50 |
| 6 | 6 | 0.84 |
| 7 | 1 | 0.14 |
| **MORTALITY** | 147 | 20.7% |
| In-hospital mortality | 147 | 20.7 |
| Out-hospital mortality | 0 | 0 |
| In-ICU mortality | 88 | 12.4 |
| Out-ICU mortality | 59 | 8.30 |
| Death > 45 days | 2 | 0.28 |

|  | *N* | *%* |
| --- | --- | --- |
| **COMORBIDITIES listed below in descending order of occurrence** (Ex. Hypertension ranked Top 1 with 48.4%) |  |  |
| **Arterial hypertension** | **344** | **48.4** |
| **BMI (Mean ± SEM)** | 27.6 ± 0.63 |  |
| >30 (Obese) | **301** | **42.3** |
| >25 and < 30 (Overweight) | 3 | 0.4 |
| **Diabetes mellitus** | **226 (type II)** | **31.6** |
| **Heart and vascular diseases** | **141** | **19.8** |
| Heart failure | 86 | 12.1 |
| Cardiomyopathies | 12 | 1.69 |
| Coronary artery diseases (CAD) | 0 | 0 |
| Atherosclerosis | 0 | 0 |
| Venous Thrombosis | 0 | 0 |
| Stroke | 0 | 0% |
| Open Heart | 31 | 4.36 |
| Previous angioplasty without placement of stent | 35 | 4.92 |
| Previous angioplasty with placement of stent | 0 | 0 |
| Others (Arrhythmia) | 7 | 0.98 |
| **Smoking behavior** | **130** | **18.3** |
| **Hyperlipidemia** | **129** | **18.1** |
| **Chronic Kidney Diseases** | **44** | **6.19** |
| Hemodialysis | 30 | 4.22 |
| Kidney transplant | 0 | 0 |
| Other (Severe chronic renal failure) | 13 | 1.83 |
| **Neurological Diseases** | **44** | **6.19** |
| Dementia | 9 | 1.27 |
| Hemiplegia | 12 | 1.69 |
| Epilepsy | 7 | 0.98 |
| Alzheimer | 8 | 1.13 |
| Parkinson | 2 | 0.18 |
| Cerebrovascular accident | 11 | 1.55 |
| **Cancer (solid tumors)** | **22** | **3.09** |
| Active cancer (AC) | 16 | 2.25 |
| History of cancer (HOC) | 6 | 0.84 |
| **Chronic Pulmonary Diseases** | **19** | **2.67** |
| Pulmonary fibrosis | 1 | 0.14 |
| Chronic obstructive pulmonary disease (COPD) | 9 | 1.27 |
| Asthma | 12 | 1.69 |
| Interstitial lung disease (ILD) | 0 | 0 |
| Cystic fibrosis | 0 | 0 |
| Atelectasis | 0 | 0 |
| Pneumonia | 2 | 0.28 |
| Lobectomy | 1 | 0.14 |
| **Blood disorders** | **4** | **0.56** |
| Hemoglobin blood disorders, *sickle cell disease (SCD)* | 0 | 0 |
| Hemoglobin blood disorders, thalassemia | 4 | 0.56 |
| Leukemia / lymphoma | 0 | 0 |
| **Liver Diseases (Cirrhosis)** | **3** | **0.42** |
| **Immuno-compromised state** | **3** | **0.42** |
| Immunodeficiency | 1 | 0.14 |
| Autoimmune diseases | 2 | 0.28 |
| **Infection** | **2** | **0.28** |
| HIV | 0 | 0 |
| HCV/HBV | 2 | 0.28 |
| **Chromosomal abnormalities** | **1 (Down syndrome)** | **0.1** |

### 3.3 Distribution of the population according to WHO Ordinal Severity Scale (WOSS)

Patients were divided into **4 groups** according to WHO Ordinal Severity Scale (**WOSS**)

“**WOSS-3**”: 130 hospitalized subjects (18.28 % of the total population) who didn’t need oxygen therapy.

“**WOSS-4**”: 393 hospitalized patients (55.27 % of the total population) who needed oxygen therapy by mask or traditional nasal cannula.

“**WOSS-5**”:153 hospitalized patients (21.52% of the total population) who needed oxygen therapy NIMV or HFNC.

“**WOSS-6**”:35 hospitalized patients (4.92 % of the total population) who needed oxygen therapy IMV & intubation.

**Table 3:** Distribution of the studied population for oxygen therapy needs related to disease progression.

| Incidence |  | *n*<sup>(1)</sup> | %<sup>(2)</sup> |
| --- | --- | --- | --- |
| **OXYGEN THERAPY (OT)** | (-) | 130 | 18.28 |
|  | (+) | 581 | 81.72 |
| **WHO Ordinal Severity Scale (WOSS)** |  |  |  |
| 0 | No clinical or virological evidence of infection | 0 | 0 |
| 1 | Ambulatory, no activity limitation | 0 | 0 |
| 2 | Ambulatory, activity limitation | 0 | 0 |
| 3 | Hospitalized, no oxygen therapy | 130 | 18.28 |
| 4 | Hospitalized, oxygen therapy by mask or nasal cannula | 393 | 55.27 |
| 5 | Hospitalized, oxygen therapy NIMV or HFNC | 153 | 21.52 |
| 6 | Hospitalized, oxygen therapy IMV and intubation | 35 | 4.92 |
| 7 | Hospitalized, IMV + additional support such as ECMO | 0 | 0 |
| 8 | Death | 147 | 20.68 |

<sup>(1)</sup>Total number of patients = 711; <sup>(2)</sup>Percentage of the total population; *Abbreviations*: OT, Oxygen Therapy; OTN, Oxygen Therapy Needs; NIMV, non-invasive mechanical ventilation; HFNC, high-flow nasal cannula; IMV, invasive mechanical ventilation.

### 3.4 Primary clinical endpoints relative to WOSS distribution in COVID-19 patients

Based on table 4, we noticed that all the deceased patients died in the hospital. There was 0% mortality in the WOSS-3 group, only 3 out of 130 (2.3%) were admitted to the ICU. While in the WOSS-4 group, many patients died (48 out of 393 (12.2%)), 1.8% were admitted to the ICU, 0.3% died in the ICU. These percentages kept on increasing significantly (P˂.0001) as the severity of the illness increased; in the WOSS-5 & 6 these percentages were respectively (43.1% – 94.3% mortality), (73.2% - 97.1% admission to ICU) & (35.9% - 91.4% mortality in the ICU). Patients who belong to WOSS-6 group (intubated) had the highest percentage of death among these groups. Same for the length of stay in the 7hospital (LOS) of total patients, it increased significantly (P˂.0001) as the severity increased. WOSS-5, 6 groups showed a higher LOS mean (12.5 days - 13.4 days respectively) than that in less severe cases.

**Table 4:** Primary clinical endpoints relative to WHO Ordinal Severity Scale (WOSS) distribution in COVID-19 patients.

|  | WOSS-3 | WOSS-4 | WOSS-5 | WOSS-6 | ANOVA |
| --- | --- | --- | --- | --- | --- |
| *n*<sup>(1)</sup> | 130 | 393 | 153 | 35 | *P* |
| **Primary Clinical Endpoints** |  |  |  |  |  |
| SURVIVAL (*n*) | 130 | 345 | 87 | 2 |  |
| MORTALITY (*n*<sup>(2)</sup>, (%<sup>(3)</sup>)) | 0 (0%) | 48 (12.2%) | 66 (43.1%) | 33 (94.3%) | <.0001 |
| In-hospital mortality | 0 (0%) | 48 (12.2%) | 66 (43.1%) | 33 (94.3%) | <.0001 |
| Out-hospital mortality | 0 (0%) | 0 (0%) | 0 (0%) | 0 (0%) | - |
| ICU admission | **3 (2.3%)** | **7 (1.8%)** | **112 (73.2%)** | **34 (97.1%)** | <.0001 |
| In-ICU mortality | **0 (0%)** | **1 (0.3%)** | **55 (35.9%)** | **32 (91.4%)** | <.0001 |
| Out-ICU mortality | 0 (0%) | 47 (12.0%) | 11 (7.2%) | 1 (2.9%) | 0.0001 |
| Death > 45 days | 0 (0%) | 0 (0%) | 1 (0.7%) | 0 (0%) |  |
| LOS of total patients (d., Mean ± SEM) | 4.9 ± 0.33 | 6.8 ± 0.18 | 12.5 ± 0.96 | 13.4 ± 1.78 | <.0001 |
| LOS of survived patients | 4.9 ± 0.33 | 6.7 ± 0.17 | 13.6 ± 1.27 | 13.5 ± 5.03 |  |
| LOS of deceased patients | - | 6.5 ± 0.68 | 11.2 ± 1.43 | 13.4 ± 1.86 |  |

<sup>(1)</sup> Total number of patients in the whole population = 711, and representative distribution per group; <sup>(2)</sup> number of deceased patients per group; <sup>(3)</sup> Percentage of deceased patients per group. *Designation:* WOSS-3, Hospitalized, no oxygen therapy; WOSS-4, Hospitalized, oxygen therapy by mask or nasal cannula; WOSS-5, oxygen therapy NIMV or HFNC; WOSS-6, oxygen therapy IMV and intubation; WOSS-8, Death (Whole Population). Significance of the observed variation was assessed by ANOVA; **F-values** determined between WOSS groups were: Mortality (101.3), LOS (44.3), In-hospital mortality (101.3), ICU admission (467.9), In-ICU mortality (230.6), Out-ICU mortality (6.93). **Abbreviations:** LOS, length of stay; d., days.

Kaplan-Meier curves (figure 1) of the combined ordinal scale of COVID-19 severity for Mortality outcome showed that there is a very significant (48.97, *P*<.0001) increase in the mortality occurrence with the increase of the illness severity; WOSS-6 group showed the highest mortality percentage in comparison with the other groups, and a LOS smaller than that in WOSS-5 group indicating that intubated patients will have a faster death. While WOSS-3 group (oxygen therapy not needed) showed 0 % mortality and a small LOS indicating a faster recovery.

**Figure 1.**
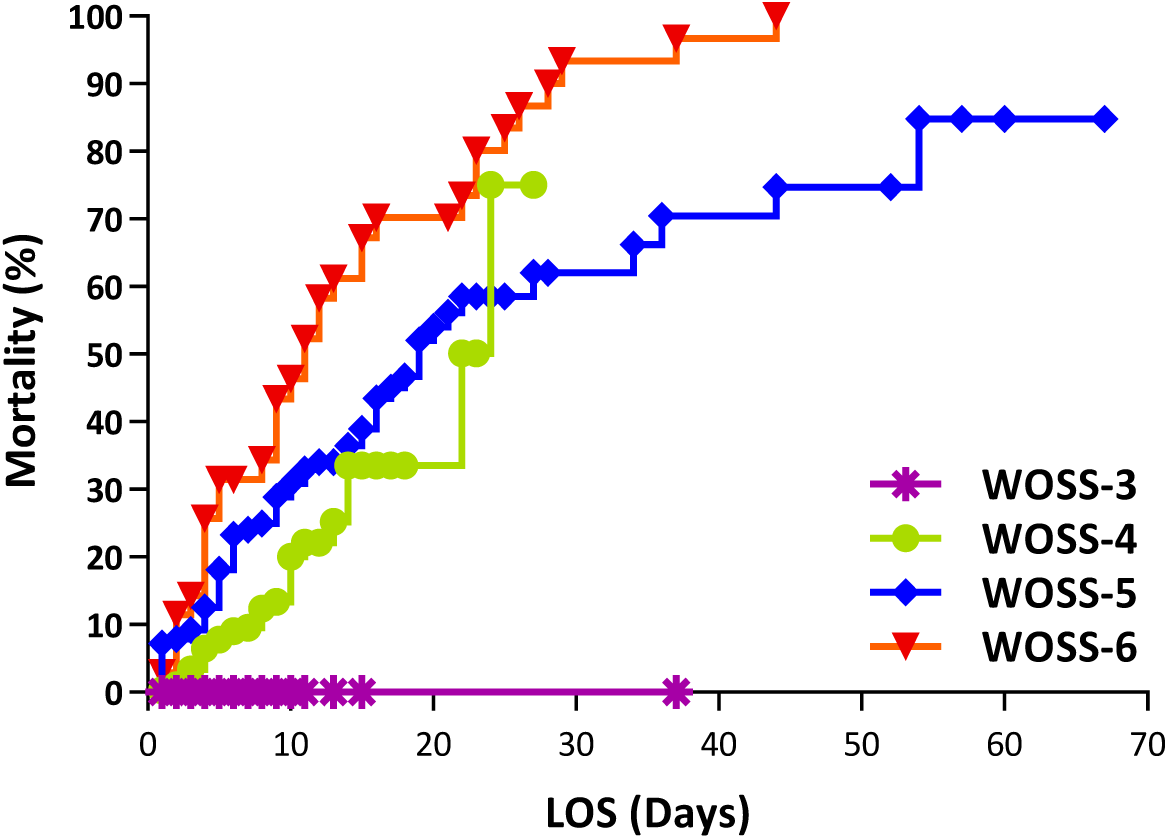
Kaplan-Meier curves of the combined ordinal scale of COVID-19 severity for Mortality outcome. Results are expressed as mortality relative to LOS (length of stay in-hospital, time from admission to death). Log-rank test for the 4 groups was: **48.97, *P*<.**0001.

### 3.5 Primary risk factors relative to WOSS distribution in COVID-19 patients

Referring to table 5, we determined the significant (p <.0001) primary related factors. The first one is the number of comorbidities, when this number increases, the severity of COVID-19 increases too (WOSS- 6 showed the highest n of comorbidities 2.8 ± 0.22) especially the predominant top 3 comorbidities which are hypertension (71.4% in WOSS-6 group), diabetes (51.4% in WOSS-6 group) & obesity (37.1% in WOSS-6 group), as usual the WOSS-6 group presented the highest percentages compared to the other less severe cases. The age is the second primary related factor. We found that the age of the total patients developing a more severe illness (WOSS- 5&6, 66.3 ± 1.27 yrs & 65.8 ± 2.34 yrs respectively) was significantly (p<.0001) greater than that who had less severe cases (WOSS-3 & 4 groups, 52.9 ± 1.88 & 63.6 ± 0.85 respectively).

**Table 5:** Primary risk factors relative to WOSS distribution in COVID-19 patients.

|  | WOSS-3 | WOSS-4 | WOSS-5 | WOSS-6 | ANOVA (P) |
| --- | --- | --- | --- | --- | --- |
| **Primary Risk Factors** |  |  |  |  |  |
| **COMORBIDITIES** |  |  |  |  |  |
| Total patients |  |  |  |  |  |
| n of comorbidities (Mean ± SEM) | **$1.5 \pm 0.12$** | **$2.0 \pm 0.07$** | **$2.2 \pm 0.12$** | **$2.8 \pm 0.22$** | **<.0001** |
| Predominant top 3 | Ob (33.8%) | Ob (52.9%) | HT (55.6%) | **HT (71.4%)** | **<.0001** |
|  | HT (31.5%) | HT (49.1%) | Db (37.3%) | **Db (51.4%)** | **<.005** |
|  | Db (22.3%) | Db (31.0%) | HD (24.8%) | **Ob (37.1%)** | **<.0001** |
| Survived patients |  |  |  |  |  |
| n of comorbidities (Mean ± SEM) | $1.5 \pm 0.12$ | $1.9 \pm 0.08$ | $2.1 \pm 0.16$ | $2.5 \pm 0.35$ | |
| Predominant top 3 | Ob (33.8%) | Ob (55.1%) | HT (55.2%) | HT (50.0%) |  |
|  | HT (31.5%) | HT (47.0%) | Db (31.0%) | Db (50.0%) |  |
|  | Db (22.3%) | Db (29.9%) | HD (23.0%) | Ob (50.0%) |  |
| Deceased patients |  |  |  |  |  |
| n of comorbidities (Mean ± SEM) | - | $2.4 \pm 0.22$ | $2.4 \pm 0.17$ | $2.9 \pm 0.23$ | |
| Predominant top 3 | - | HT (64.6%) | HT (56.1%) | HT (72.7%) |  |
|  |  | Db (39.6%) | Db (45.5%) | Db (51.6%) |  |
|  |  | Ob (37.5%) | HD (28.8%) | Ob (36.41%) |  |
| **Age of total patients (yrs., Mean ± SEM)** | **$52.9 \pm 1.88$** | **$63.6 \pm 0.85$** | **$66.3 \pm 1.27$** | **$65.8 \pm 2.34$** | **<.0001** |
| Age of survived patients | $52.9 \pm 1.88$ | $61.3 \pm 0.87$ | $63.9 \pm 1.75$ | $60.5 \pm 1.77$ | |
| Age of deceased patients | - | $80.1 \pm 1.63$ | $69.3 \pm 1.74$ | $66.2 \pm 2.47$ | |

**F-values** between WOSS groups: number of comorbidities (9.28), obesity (15.71), hypertension (8.75), diabetes (4.67), heart diseases (3.70), age (16.38). Ob, Obesity; HT, Hypertension; Db, Diabetes; HD, Heart and Vascular Diseases; Yrs., years.

Table 6 showed the same results using the OR. The OR concerning the mortality in case of WOSS-6 versus WOSS-3 is the greatest one (3497.4 - p˂.0001 significant) compared to that of WOSS-5 versus WOSS-3 (198.4 - p˂.0001 significant) which is also higher than the OR in case of WOSS-4 versus WOSS-3(36.6 - p˂.0001 significant) this means that severity of the disease is loads significantly positively correlated with mortality. The OR concerning the ICU admission in case of WOSS-6 versus WOSS-3 is the greatest one (1439.3 - p˂.0001 significant) compared to that of WOSS-5 versus WOSS-3 (115.6 - p˂.0001 significant) this means that severity of the illness is also very significantly positively correlated with the admission to the ICU. The OR concerning the comorbidities in case of WOSS-6 versus WOSS-3 is as well the greatest one (6.19 - p˂.0001 significant) compared to that of WOSS-5 versus WOSS-3 (2.16 - p˂.001 significant) which is in its turn higher than the OR in case of WOSS-4 versus WOSS-3(1.75 - p˂.005 significant) this means that severity of COVID-19 is significantly positively correlated with comorbidities (2 or more as a cut-off). The OR concerning the age in case of WOSS-6 versus WOSS-3 is the greatest one (3.85 - p˂.001 significant) compared to that of WOSS-5 versus WOSS-3 (2.81 - p˂.0001 significant) which is also higher than that in case of WOSS-4 versus WOSS-3(2.51 - p˂.0001 significant) this means that severity of the illness represented by the need of intubation is significantly positively correlated with age (64 years or more as a cut-off).

**Table 6:**
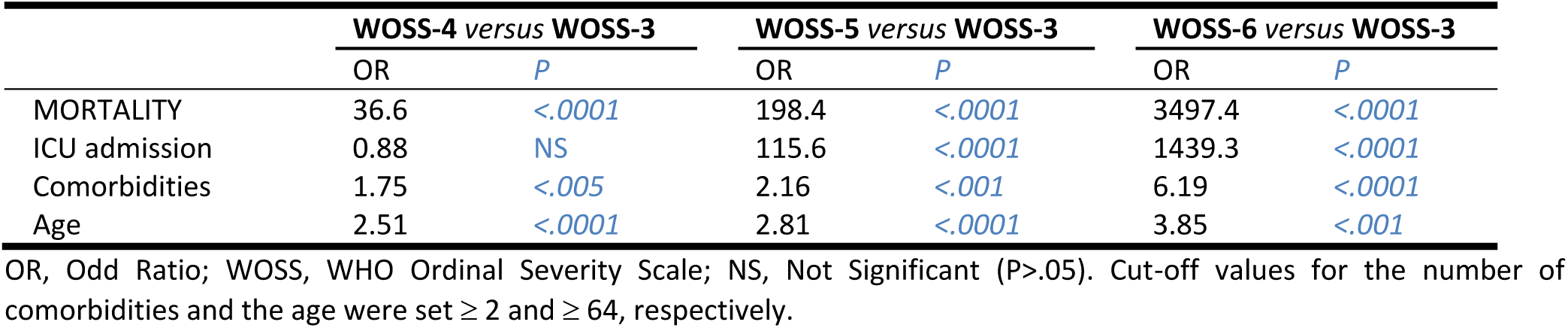
Significance variations of primary clinical endpoints in correlation to COVID-19 illness severity.

### 3.6 Evaluation of the main symptoms and the pulmonary inflammatory lesions relative to WOSS distribution in COVID-19 patients

Table 7 shows the symptoms evolution from WOSS-3 to WOSS-6. Nausea decreased significantly (p˂.01) from 9.2 till 2.9%, diarrhea decreased significantly (p ˂.005) from 13.8 till 2.9% and vomiting also decreased significantly (p ˂.05) from 13.8 till 2.9 % with the progression of the illness severity. Apnea varied significantly (p ˂.0001) between the different groups; it occurred in WOSS-4 extremely high (74.8%) probably due to the use of mask or nasal cannula while it occurred in WOSS-5 extremely low (34.6%) indicating beneficial impact of NIMV or HFNC. Fever occurrence increased from 44.6 % till 60 with the increases of severity. The percentage of pulmonary inflammatory lesions and its distribution relative to WOSS scale in COVID-19 patients. We noticed that the percentage of pulmonary inflammatory lesions increased significantly (<.0001) from 16.3 ±1.71 till 50 ± 5.72% with the increase of the illness severity (WOSS-3 till WOSS-6). Those who were intubated had the highest percentage of pulmonary inflammation. Concerning the distribution of pulmonary inflammatory lesions, we found that: intubated patients had a high significant (<.0001) occurrence of pneumothorax inflammation lesions (18.2%). Patients who belong to WOSS-5 group had the highest occurrence of pulmonary inflammatory lesions in the lower zones (15.9%), it didn’t occur in WOSS-6 group (0%). Bilateral PIL significantly (P˂.05) occurred in all patient’s groups. PIL increased with illness severity, lesions are distributed bilateral and in lower zones, a high occurrence of pneumothorax was revealed.

**Table 7:** Main symptoms and pulmonary inflammatory lesions relative to WOSS distribution in COVID-19 patients.

|  | WOSS-3 | WOSS-4 | WOSS-5 | WOSS-6 | P | F |
| --- | --- | --- | --- | --- | --- | --- |
| **n<sup>(1)</sup>** | **130** | **393** | **153** | **35** |  |  |
| **MORTALITY (n<sup>(2)</sup>, (%<sup>(3)</sup>))** | **0 (0%)** | **48 (12.2%)** | **66 (43.1%)** | **33 (94.3%)** | **<.0001** | **7.34** |
| **Main symptoms** |  |  |  |  |  |  |
| Apnea | 56 (43.1%) | 294 (74.8%) | 53 (34.6%) | 16 (45.7%) | <.0001 | 36.02 |
| Nausea | 12 (9.2%) | 35 (8.9%) | 5 (3.3%) | 1 (2.9%) | <.01 | 3.93 |
| Diarrhea | 18 (13.8%) | 43 (10.9%) | 5 (3.3%) | 1 (2.9%) | <.005 | 4.36 |
| Vomiting | 18 (13.8%) | 31 (7.9%) | 8 (5.2%) | 1 (2.9%) | <.05 | 2.93 |
| Fever | 58 (44.6%) | 206 (52.4%) | 84 (54.9%) | 21 (60.0%) | <.05 | 3.28 |
| **Pulmonary inflammatory lesions (PIL)<sup>(4)</sup>** |  |  |  |  |  |  |
| % of PIL | 16.3 ± 1.71 | 31.2 ± 0.47 | 43.5 ± 3.08 | 50.0 ± 5.72 | <.0001 | 17.53 |
| **Distribution of pulmonary inflammatory lesions (% of patients per group)<sup>(5)</sup>** |  |  |  |  |  |  |
| Bilateral | 93.0% | 87.7% | 79.5% | 63.6% | <.05 | 2.90 |
| Peripheral | 44.2% | 55.0% | 50.0% | 36.4% | NS | 0.99 |
| Multilobar | 41.9% | 49.5% | 29.5% | 18.2% | NS | 0.68 |
| Lower zones | 14.0% | 15.5% | 15.9% | 0.0% | <.05 | 3.21 |
| Pneumothorax | 0.0% | 0.0% | 0.0% | 18.2% | <.0001 | 22.45 |
| **Pulmonary Embolism (PE)** |  |  |  |  |  |  |
| % of PE | 0 | 0 | 0 | 0 | - | - |
| **Venous Thromboembolism (VTE)** |  |  |  |  |  |  |
| % of PE | 0 | 0 | 0 | 0 | - | - |

<sup>(1)</sup>Total number of patients in the whole population = 711, and representative distribution per group; <sup>(2)</sup>number of deceased patients per group; <sup>(3)</sup>Percentage of deceased patients per group; <sup>(4)</sup>Quantification of the pulmonary inflammatory lesions and their distribution were evaluated by chest CT scan at admission and expressed relative to the total surface of the lung; <sup>(5)</sup>Percentage of patients in each group with bilateral/peripheral/ multilobar/lower zones /pneumothorax PIL. *Designation:* WOSS-3, Hospitalized, no oxygen therapy; WOSS-4, Hospitalized, oxygen therapy by mask or nasal cannula; WOSS-5, oxygen therapy NIMV or HFNC; WOSS-6, oxygen therapy IMV and intubation. *Abbreviations:* PIL, pulmonary inflammatory lesions; PE, pulmonary embolism; CT, computed tomography; VTE, venous thromboembolism.

### 3.7 SARS-CoV-2 RNA and IgM/IgG antibodies variations relative to the WHO ordinal scale of COVID-19 severity

Figure 2 shows that all the patients regardless the group to which they belong had a positive SARS-COV-2 PCR test (without any significant variation). While figures B/C indicate that the *IgM* antibodies variations between the different WOSS groups & between subgroups survived vs deceased were significant (P<.001, P<.05, & P<.01 respectively). Referring to figure C, we found that deceased patients, whatever the WOSS group is, had higher SARS-CoV-2 IgM antibodies than the survived ones. SARS-CoV-2 IgM antibodies decreased significantly with the progression of the disease severity; WOSS-6 group presented the smallest level of SARS-CoV-2 IgM antibodies (about 7 in case of deceased patients & about 4.2 in case of survival). Figures D/E showed no significant (P>.05) SARS-CoV-2 IgG antibodies variations neither between the different WOSS groups nor between the subgroups: survived vs deceased.

**Figure 2.**
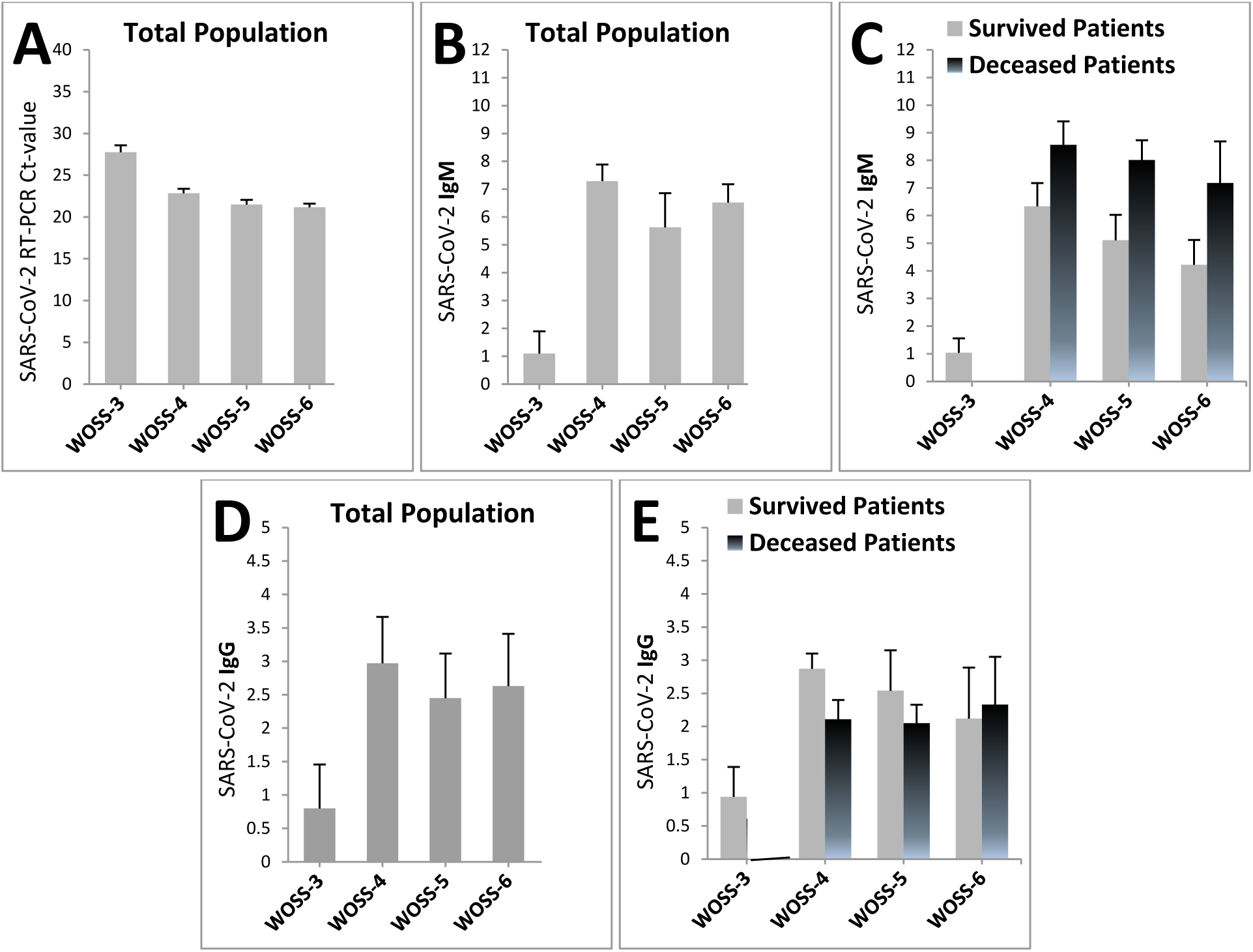
SARS-CoV-2 variations of the WHO ordinal scale of COVID-19 severity. SARS-CoV-2 RNA (A) and IgM/IgG antibodies (B-E) were determined as described in Methods. Results are expressed as the Mean ± SEM in total population (A,B,D) or distributed between survived and deceased patients (C,E) per WOSS groups. Measures (M) were determined at the first day of admission. RT-PCR Ct values were shown. Cutoffs for IgM and IgG were 1. For RNA RT-PCR Ct and IgG variations between groups: no significances were detected (P>.05). No significances were obtained between subgroups: survived vs deceased. For IgM variations between WOSS groups, significances were detected for total population, survived and deceased patients (P<.001, P<.05, and P<.01, respectively).

### 3.9 Treated Population with antivirals or immunosuppressive

Patients received corticosteroid therapy-enhanced standard care (CTSC) alone or combined with antivirals (Remdesivir or Lopinavir/Ritonavir) or the immunosuppressive anti-IL-6R agent Tocilizumab, forming three comparative arms: Control (−, n=502), Antivirals (A, n=108), and Immunosuppressive (B, n=101). Population was stratified by WHO Ordinal Severity Scale (WOSS), yielding 11 subgroups (WOSS-3 through WOSS-6), with no WOSS-3 patients receiving treatment (B).Before subgroup division, mortality was comparable across groups (20.9%, 19.4%, and 18.8% for −, A, and B respectively), while LOS differed significantly, with (B) longest and (−) shortest. After stratification, WOSS-4 showed the most notable benefit: (B) achieved the lowest mortality (1.8%, p<0.01) and significantly longer LOS versus (−). WOSS-5 also showed significant mortality reduction, with (B) again more efficient (26.4%), and both (A) and (B) extending LOS considerably. In WOSS-6, neither treatment significantly reduced mortality — all patients receiving CTSC alone died — though both (A) and (B) extended LOS relative to (−), reflecting prolonged survival rather than recovery. No deaths occurred in WOSS-3.OR analysis confirmed no significant mortality difference between (A) and (−) across all WOSS groups, while (B) consistently outperformed (−) in WOSS-4, 5, and 6 (OR=0.11, 0.38, and 0.08 respectively), and surpassed (A) significantly in WOSS-4 (OR=0.12, p<0.05). These findings underscore the relevance of Tocilizumab, particularly in moderate illness, while highlighting its limited efficacy in severe cases.

**Table 8:**
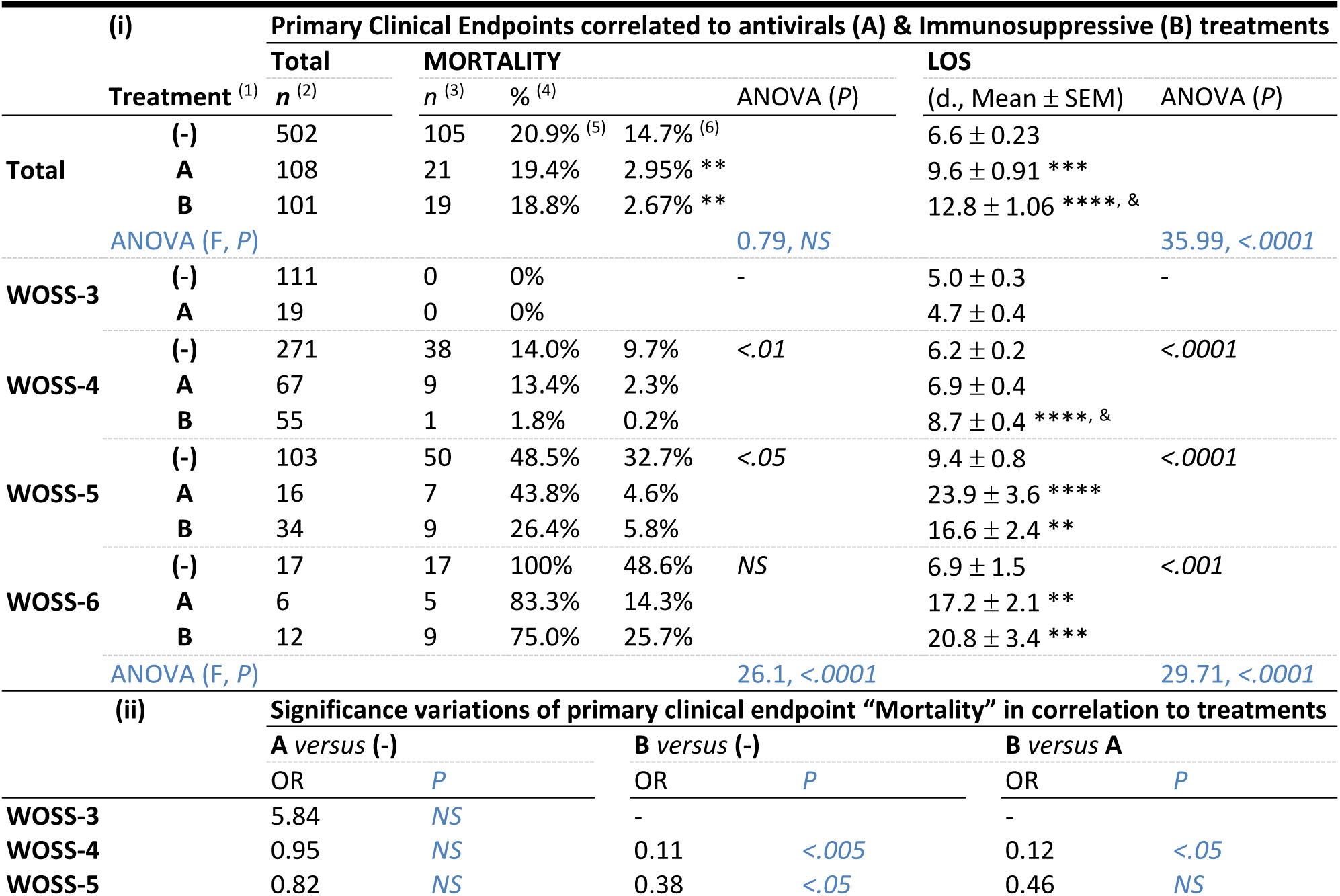
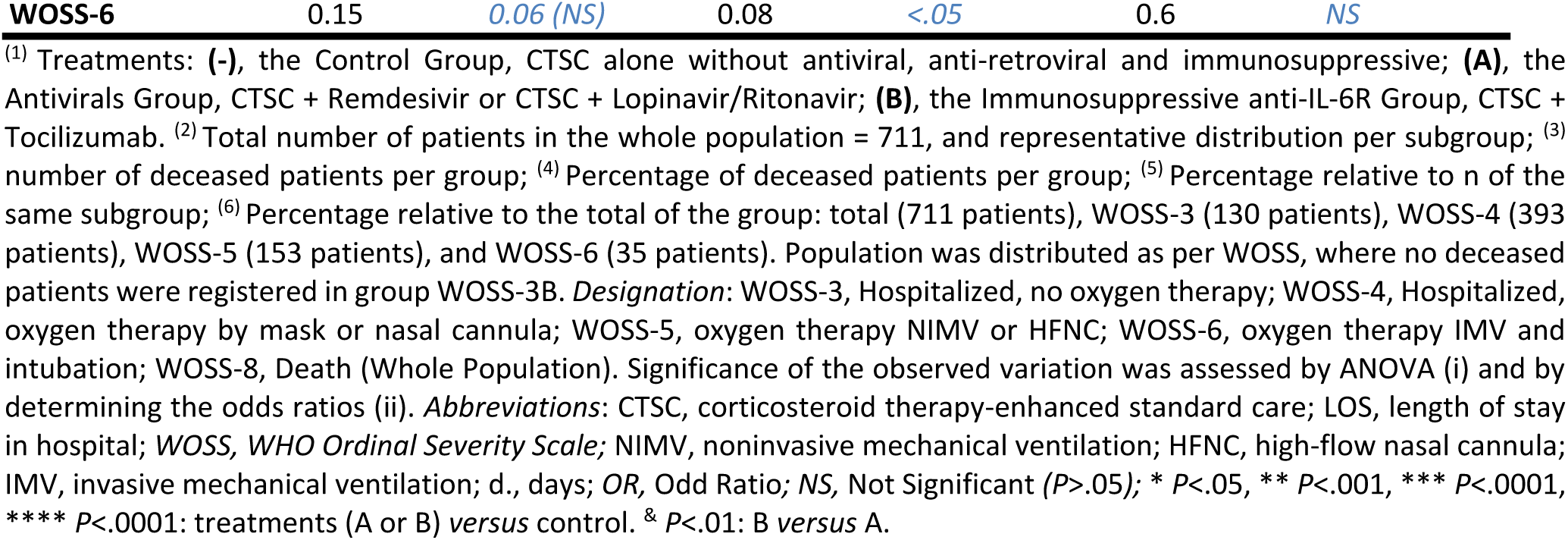
Primary clinical endpoints relative to WOSS distribution in COVID-19 treated patients.

Referring to the Kaplan-Meier curves of the combined ordinal scale of COVID-19 severity for Mortality outcome showing the impact of treatments, we noticed that in WOSS-6 group (intubated), there is approximately the same high mortality % whatever the treatment is (-),(A), or (B). LOS in the woss-6(B) subgroup was the longest while in woss-6(-) subgroup it was the lowest indicating that (-) treatment leads to the fastest death. In WOSS-5 group, we noticed that treatment (-) led to the highest mortality %. The lowest mortality % was in the woss-5 (B) subgroup. This means that (B) reduces the mortality. (B) & (A) showed a little bit difference in the mortality % (almost the same). In case of (B) treatment, we found that patients stayed the longest time in the hospital in comparison with all the other cases. (-) leads to the fastest death in WOSS-5 group. In WOSS-4 group: we noticed that group (-) had the highest mortality % as usual. This treatment is not efficient as the others. (A) is better than (-) since the results showed a lower death %. Group (B) showed the lowest mortality %. WOSS-4 (B) showed the smallest LOS this means that (B) reduces well the mortality and it is considered as the best choice to a faster efficient recovery in a non-very severe case. Ιn WOSS-3 group, there is 0 % mortality in both cases. WOSS-3 (A) subgroup showed the smallest LOS indicating a faster recovery. All of these results are significant (99.06, *P*<.0001).

**Figure 3.**
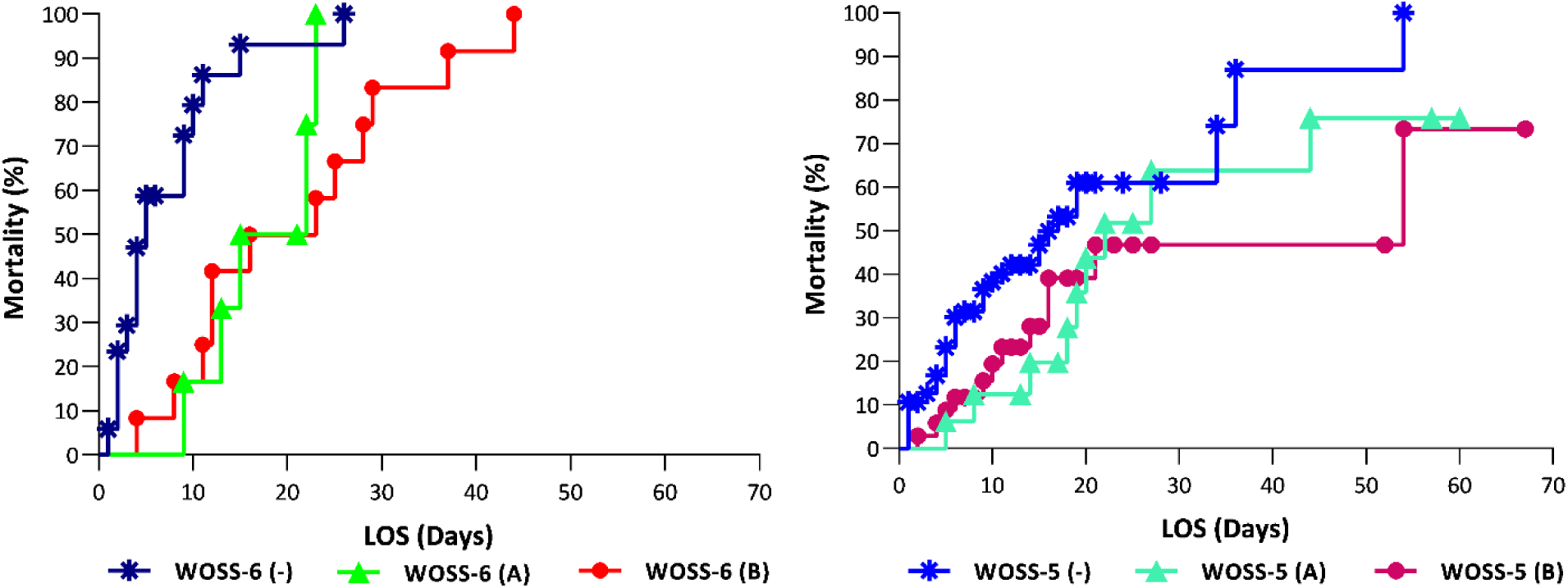
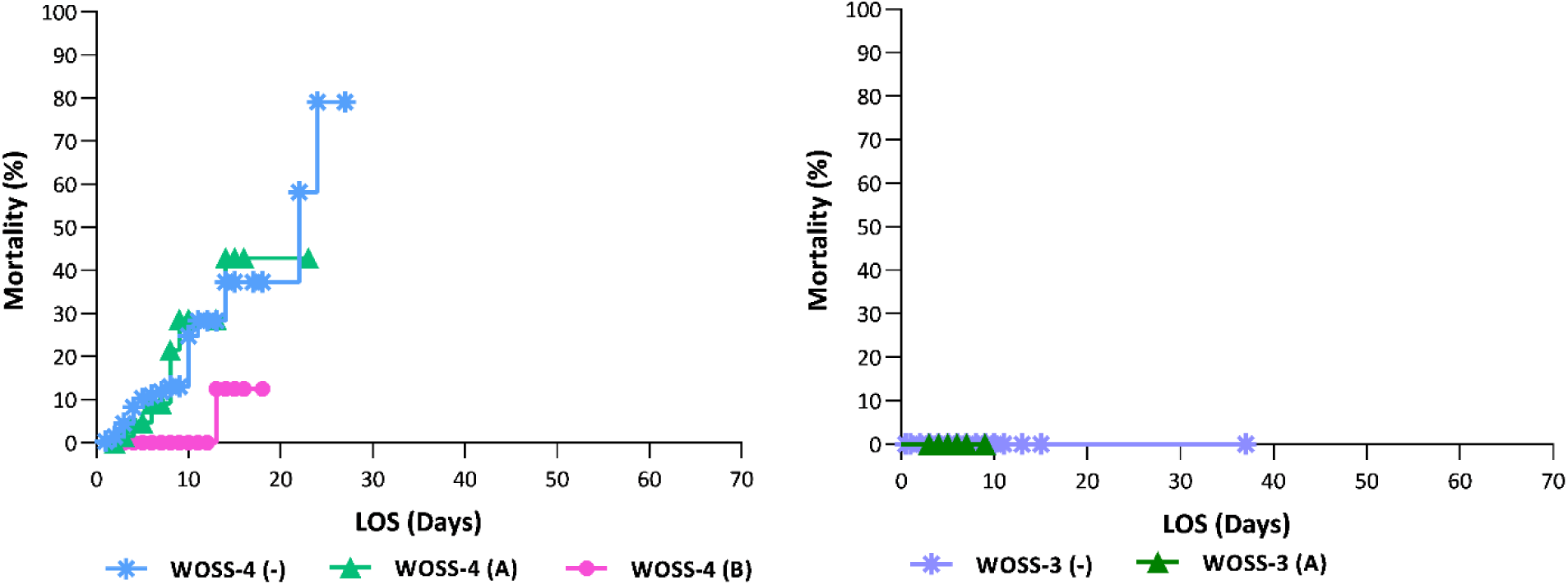
Kaplan-Meier curves of the combined ordinal scale of COVID-19 severity for Mortality outcome: impact of treatments. 3 comparative arms were studied: control (without antivirals, anti-retrovirals and immunosuppressive) designed by Groups (-), antivirals (Remdesivir, Lopinavir/Ritonavir) designed by Groups (A), and immunosuppressive anti-IL-6R (Tocilizumab) designed by Groups (B). Population was distributed as per WOSS, where no deceased patients were registered in group WOSS-3 (B). Results are expressed as mortality relative to LOS (length of stay in-hospital, time from admission to death). Log-rank test for the 11 comparative subgroups**: 99.06, *P*<.**0001.

Analysis of Table 10 revealed that while no significant differences existed between treatment groups (−), (A), and (B) in the overall distribution of main symptoms and pulmonary inflammatory lesions (PIL) prior to subgroup stratification, significant variations emerged following WOSS-based distribution. Treatment (B) proved most effective overall: it produced the lowest rates of fever in WOSS-4 and WOSS-5 subgroups (58.2% and 58.8% respectively) and the greatest reduction in apnea in WOSS-5 and WOSS-6 (29.4% and 58.3%), while treatment (A) outperformed (−) in WOSS-3 for both symptoms. Regarding pulmonary involvement, (B) achieved the greatest reduction in PIL — from 52.2% in (−) to 32.9% in (A) and 26.9% in (B) (p<0.0001) — with the lowest PIL percentages in WOSS-4 and WOSS-6 subgroups (22.4% and 45.2%), and the lowest pneumothorax occurrence in WOSS-6 (16%). No significant inter-group differences were observed for nausea, diarrhea, vomiting, or bilateral and lower-zone lung distribution. In conclusion, treatment (B) was the most effective in reducing fever, apnea, and pulmonary inflammation, and (A) showed improvement over standard care (−), though residual inflammation remained substantial across all groups.

**Table 9:**
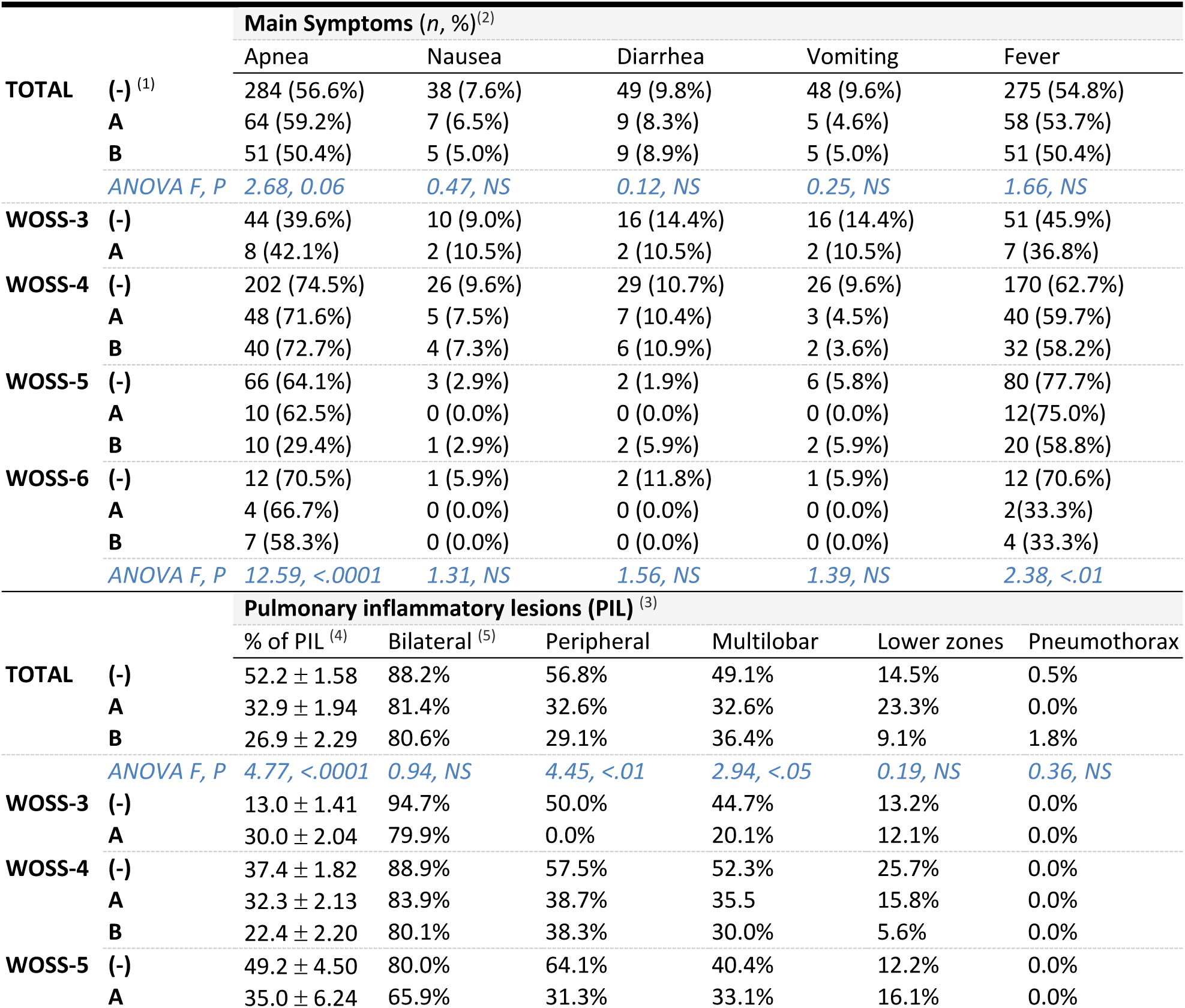
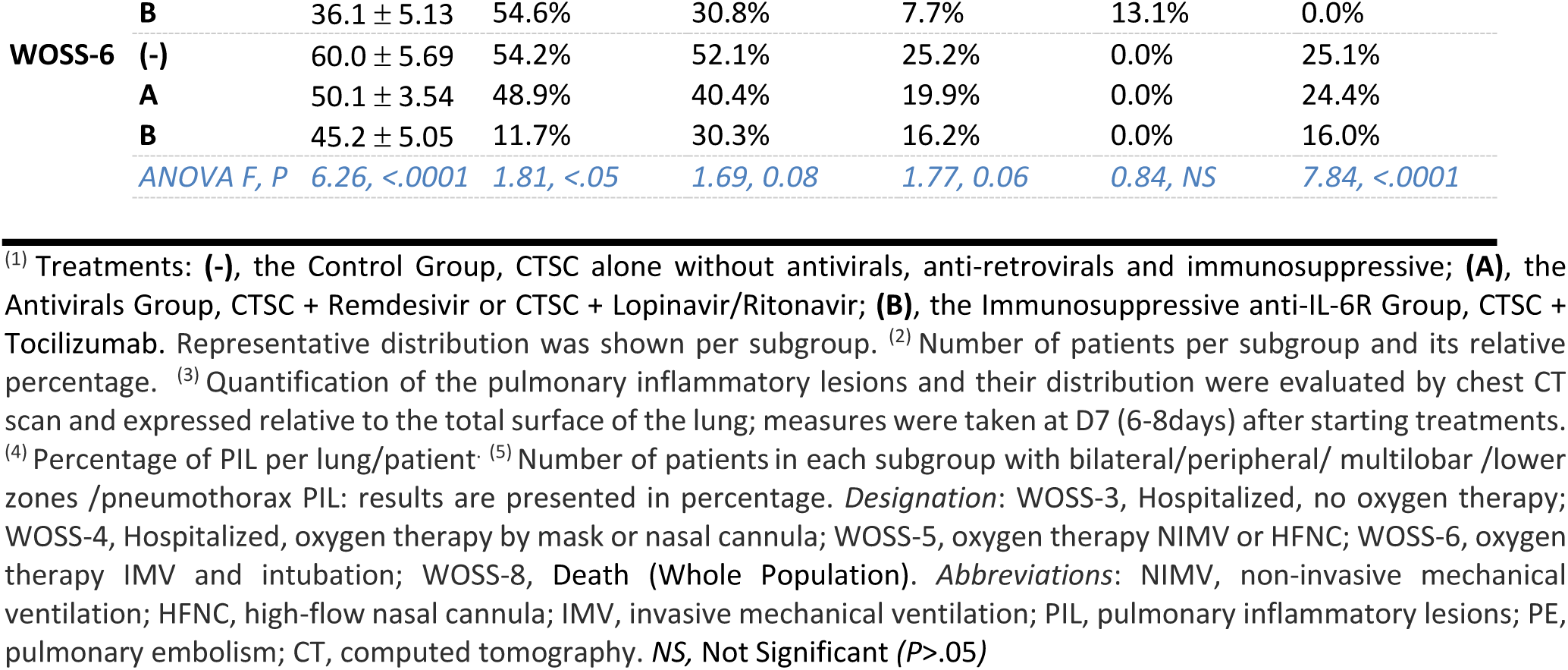
Main symptoms and pulmonary inflammatory lesions (PILs) correlated to antivirals, anti-retrovirals (A) and Immunosuppressive (B) treatments.

### 3.10 Laboratory biomarkers patterns of responses in COVID-19 treated patients

Figure 4 showed that the LDH level (the normal range is 135-225 U/L) in WOSS- 6 groups was significantly (P˂.0001) reduced from 850 (-) to 627 & 352.4 U/L when the patients were treated with A & B respectively. (B) is more efficient than (A) but we noticed that the LDH level is still higher than the normal range even with (B) in the severe stages of the illness. Same for CRP level (normal range is 0-6 mg/L), it was significantly (P˂.0001) reduced in WOSS- 6 groups from 139.5 (-) to 108 & 80.1 mg/L when the patients were treated with A & B respectively. (B) is more efficient than (A) but we noticed that the CRP level is still higher than the normal range even with (B) in the severe stages of the illness.

**Figure 4:**
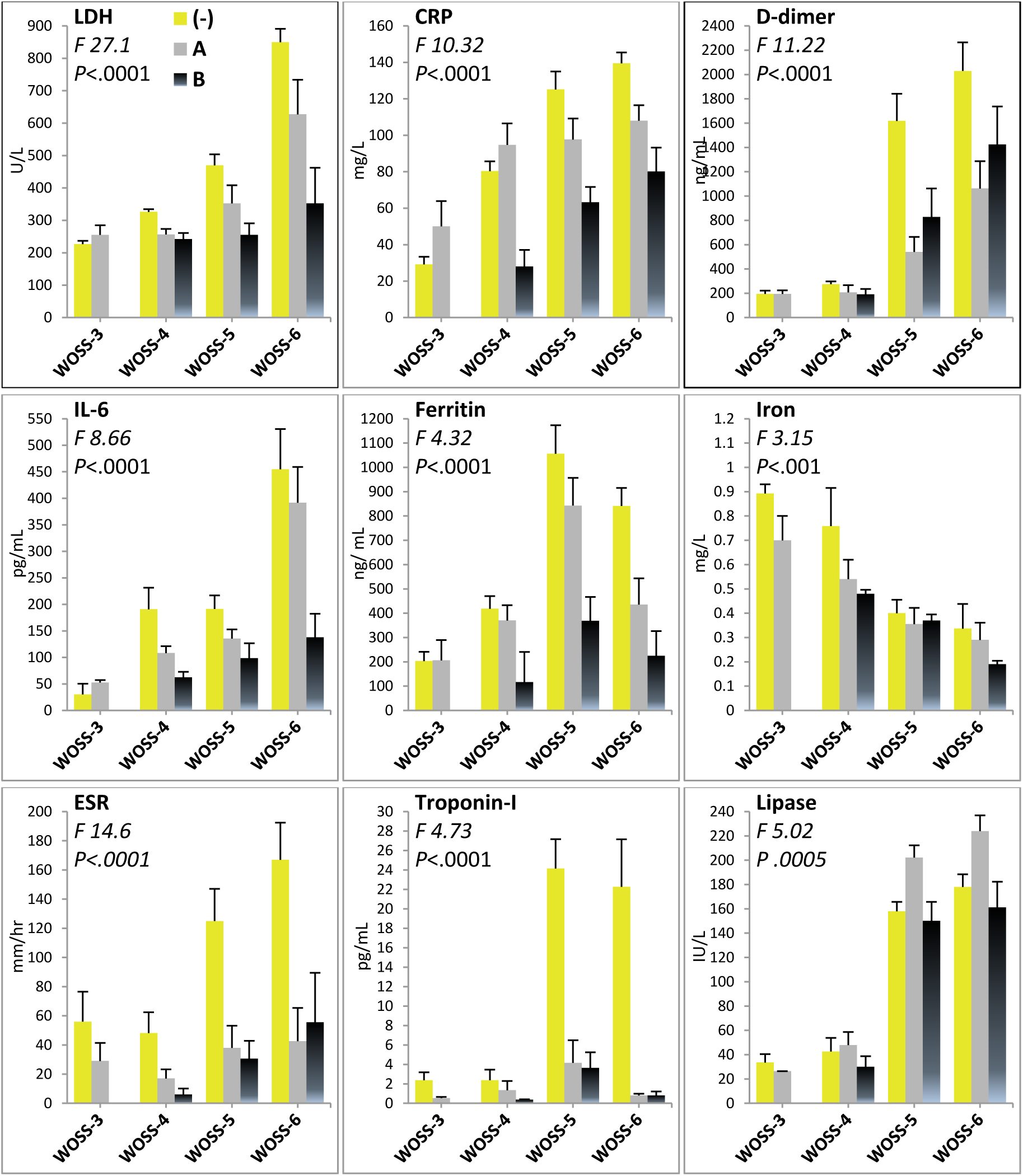
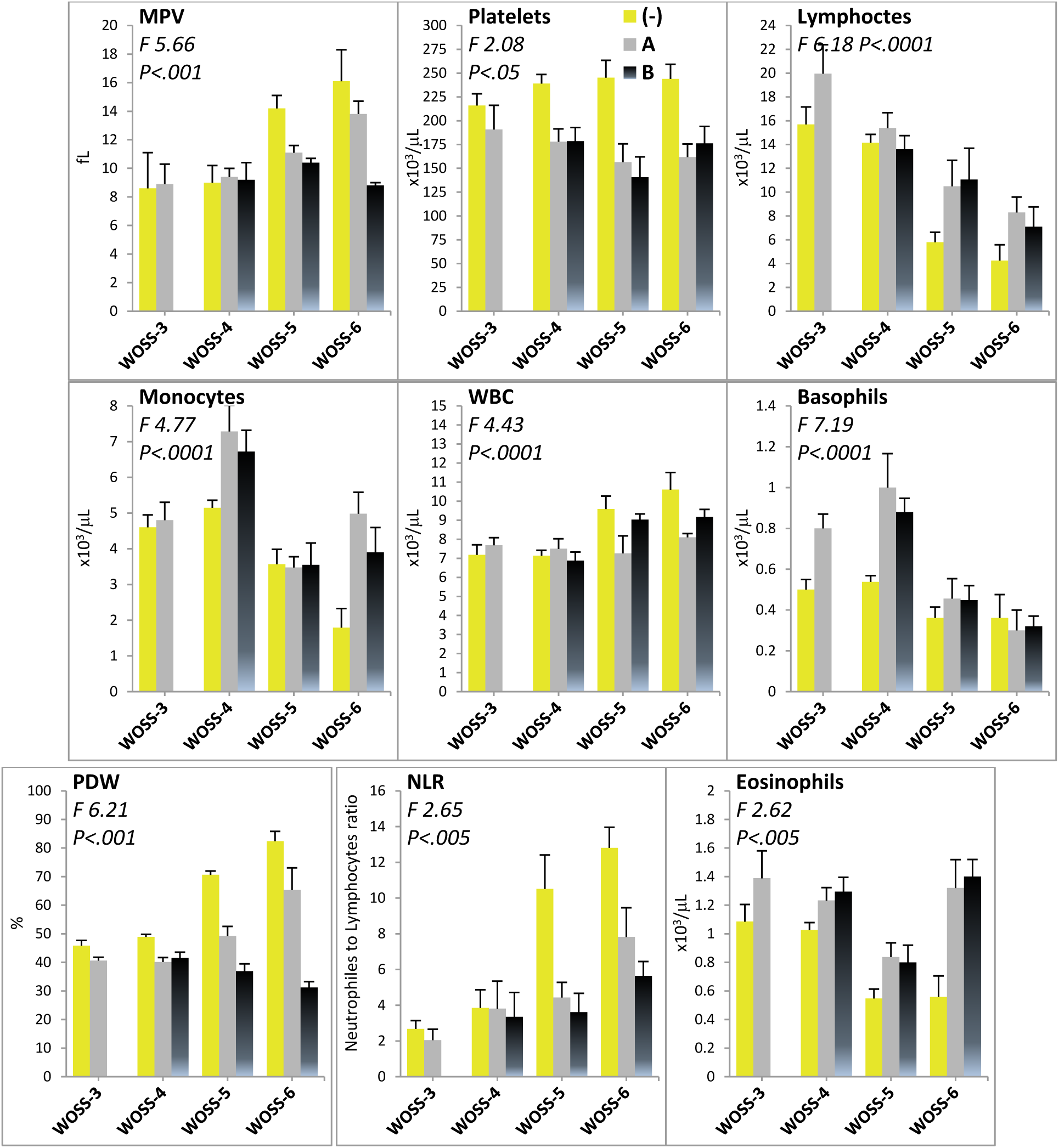
Laboratory biomarkers patterns of responses in COVID-19 treated patients. Patients were treated as indicated in methods with CTSC with or without antivirals (Lopinavir/Ritonavir, Remdesivir) or immunosuppressive anti-IL-6R (Tocilizumab); 3 groups were studied: (-), the Control Group, CTSC alone without antivirals and immunosuppressive; (A), the Antivirals Group, CTSC + Remdesivir or CTSC + Lopinavir/Ritonavir; (B), the Immunosuppressive anti-IL-6R Group, CTSC + Tocilizumab. Results expressed in absolute values are presented as means ± SEM. Two measures were taken: the first at time of receiving treatments and the second after 7 days (D6-D8) of receiving treatments, and the mean of the obtained values for the 2 measures was presented. Significance of the observed variation was assessed by ANOVA: the F-statistic, calculated as the variation between the sample means divided by variation within the samples and representing the degree of dispersion among different WOSS groups, was used to quantify the models A versus B versus (-). Only biomarkers that showed significant variations are presented.

The normal range of the D-dimer is 0-500 ng/mg. The D-dimer level was significantly (P˂.0001) reduced in WOSS- 6 group from 2030 (-) to 1062.4 (half) & 1424.3 mg/L when the patients were treated with A & B respectively. (A) is more efficient than (B) but we noticed that the CRP level is still higher than the normal range even with (A) &(B) in the severe stages of the illness. The normal range of the IL-6 is 0-43.5 pg /mL. The IL-6 level was significantly (P˂.0001) reduced in WOSS- 6 group from 454.8 (-) to 391.8 & 137.9 when the patients were treated with A & B respectively. (B) is indeed more efficient than (A) but we noticed that the IL-6 level is still higher than the normal range even with (B) in the severe stages of the illness. Same for ferritin (normal range is 15-150 ng /mL), its level was significantly (P˂.0001) reduced in WOSS- 6 group from 841.7 (-) to 435.4 & 224.8 when the patients were treated with A & B respectively. (B) is more efficient than (A) but we noticed that the ferritin level is still higher than the normal range even with (B) in the severe stages of the illness. Same for the neutrophil count (normal range is 2-7*10^3^ /μL), its level was significantly (P˂.0001) reduced in WOSS- 6 group from 54.6 (-) to 48.3 & 40.1 when the patients were treated with A & B respectively. (B) is more efficient than (A) but we noticed that the neutrophil count is still higher than the normal range even with (B) in the severe stages of the illness. The lymphocyte count (normal range is 1.5-4*10^3^ /μL, its level was significantly (P˂.0001) increased in WOSS- 6 group from 4.3 (-) to 8.3 & 7.1 when the patients were treated with A & B respectively. (A) is more efficient than (B) in the severe stage of the illness. The normal range of troponin-I is ˂15 pg /mL. The troponin-I level was significantly (P˂.0001) reduced in WOSS- 6 group from 22.3 (-) to 0.8 when the patients were treated with A & B. Both (B) & (A) are incredibly efficient; troponin-I became normal after treatment. The normal range of NLR is 0.78 – 3.53. The NLR significantly (P˂.005) decreased in WOSS- 6 group from 12.8 (-) to 7.8 & 5.6 when the patients were treated with A & B respectively. (B) is the most efficient but not efficient as proved previously.

## 4 Discussion

In the first part of this study, we explored the potential association between mortality and severity of COVID-19 illness. We started by classifying the population (n=711) according to the WHO ordinal severity scale. We divided the population into 4 groups (WOSS-3 till WOSS-6) having an increased illness severity related to the oxygen therapy. WOSS-6 group, including all the intubated patients developing critical illness, had the highest occurrence of mortality, the highest admission to and mortality in the ICU indicating a significant positive correlation between severity of the illness and mortality. These results didn’t surprise us since their blood tests showed the highest levels of inflammatory biomarkers such as LDH, D-dimer, IL-6, CRP, ferritin, NLR and the lowest lymphocyte count (lymphopenia). Markers of adaptive immunity such as lymphocytes decreased significantly leading to a marked rise of the NLR ratio. Our results are in accordance with so many international scores were it was mentioned that D-dimer, LDH, LC, CRP, NLR, & IL-6, among other parameters to be considered, are indeed related to the development of a critical illness after being infected with SARS-CoV-2.The rise in the comorbidities number was tightly associated to mortality in the studied population. In fact, mortality had the highest occurrence in severe cases (woss-6 group) that had the greater number of comorbidities especially the arterial hypertension that may lead to more complications (it’s important to state that woss-3 group that didn’t need oxygen therapy suffered from less comorbidities mainly of obesity not hypertension), diabetes and obesity indicating that these data come in agreement with the fact that 5 out of 18 scores, mentioned in previous studies, stated the importance of the number of comorbidities while calculating the points that is needed to identify if a patient have a low, intermediate or high risk of mortality. Same for the age of the patient, this was the second significant risk factor positively correlated with the illness severity. When the age increases, so does the severity and consequently the mortality. WOSS-6 including elder patients showed the highest mortality percentage. These data also come in agreement with 10 scores out of 18 that insisted on the significant link between age and the risk of developing severe illness leading to death. WOSS-3 group showed the smallest LOS among the different groups, it’s logical since patients didn’t need oxygen therapy, they had smaller levels of inflammatory markers therefore they had no severe illness and consequently will heal and recover faster leading to a decline in the duration of stay in the hospital. Patients who belong to WOSS-6 group stayed for a short time in the hospital highlighting a faster death probably because of COVID-19’s lung damage, COVID-19 can cause lasting lung damage. COVID-19 can cause lung complications such as pneumonia and, in the most severe cases, acute respiratory distress syndrome, or ARDS. Sepsis, another possible complication of COVID-19, can also cause lasting harm to the lungs and other organs. Newer coronavirus variants may also cause more airway disease, such as bronchitis, that may be severe enough for hospitalization. As we have learned more about SARS-CoV-2 and resulting COVID-19, we have discovered that in severe COVID-19, a significant pro-inflammatory condition can result in several critical diseases, complications and syndromes, if COVID-19 pneumonia progresses, more of the air sacs can become filled with fluid leaking from the tiny blood vessels in the lungs. Eventually, shortness of breath sets in, and can lead to acute respiratory distress syndrome (ARDS), a form of lung failure. Patients with ARDS are often unable to breathe on their own and may require ventilator support to help circulate oxygen in the body. [9] The mortality rise was significantly linked with a higher percentage of pulmonary inflammatory lesions; pneumothorax inflammation was shown only in case of WOSS-6 group probably because patients are already receiving mechanical ventilation. [10] Pneumothorax is the medical term for a collapsed lung. Air enters the pleural space. This can happen when an open injury in a lung tissue causes air to leak into the pleural space. The resulting increased pressure on the outside of your lung causes it to collapse, pneumothorax results from an injury to the lungs. It has been suggested that the cytokine storm contributed to the development of ARDS (acute respiratory distress syndrome) in COVID-19 patients due to the high levels of IL-6 mainly. Compared to survivors, patients who died within 28 days had increased levels of IL-6 [11] Those who had less inflammatory markers had better survival rate. The high inflammation (caused by prostaglandins and IL-6) can justify the elevation in the woss-6 patient’s body’s temperature. Prominent levels of anti SARS-CoV-2 antibodies are associated with severe diseases [12] immune system is activated to clear the coronaviruses, our study showed the same results, WOSS-4, 5&6 had greater levels of IgM compared to WOSS-3 group. Those who belonged to WOSS-6 group (severe case) have shown significantly lower IgM SARS-CoV-2 antibodies compared to those with less illness severity; this can be probably justified by the fact that extremely high IL-6 levels produced in severe cases can lead to an exhaustion in lymphocytes B producers of the anti-SARS-CoV-2 antibodies. Woss-3 patients didn’t have IgG (less than 1), low level of IgM antibodies reflecting a low viral load were only detected this can be justified by the fact that those patients were recently infected about 14 to 21 days before their entrance to the hospital while the other groups had a bigger viral load consequently a higher IgM antibodies amount and IgG were detected indicating a non-recent infection. Surviving patient’s subgroup showed lower levels of these antibodies than in deceased ones, they should have a smaller viral load promoting survival.

Concerning the treatment, three comparative arms were studied: the Control Group: CTSC alone, designed by (-), Antivirals or anti-retrovirals Group: CTSC + Remdesivir or Lopinavir/Ritonavir, designed by (A) and Immunosuppressive anti-IL-6R Group: CTSC + Tocilizumab, designed by (B). Our objective was to explore which one is the most efficient especially to treat severe cases thus reducing the mortality rate, which was high in severe cases as shown before. To our knowledge, no studies have yet focused on that correlation. The occurrence of mortality wasn’t significantly reduced in WOSS-6 group which is the most severe case regardless the type of treatment; WOSS-6 (B) subgroup showed 75 % death a really high percentage even if it is less than that in case of WOSS-6 (-) subgroup (100% mortality). This can be justified by the lung injuries that can be hardly healed and the very high IL-6 level even if tocilizumab decreased it but it’s still a high level greater than the normal range. The results also showed in this subgroup a reduction in other inflammatory biomarkers as developed in detail in the results part but the levels after the treatment are still higher than the normal range; the cytokine storm main crucial cause of death is still present causing damages to the lung worsening the patient’s status; treatment (B) is partially and not really efficient against severe COVID-19 cases. We should evaluate the impact of other treatments to reduce more and more the inflammation since tocilizumab is specific for IL-6 it reduces it but there are many others elevated inflammatory biomarkers not affected by Actemra. We can focus in a new study on new parameters like PAF(platelet-activating factor) a powerful mediator of inflammation and leukotriene and prostaglandin.

No significant variation between (-) and (A) has been revealed, this means that antivirals and anti-retrovirals are not efficient as a treatment against this disease. No clear molecular mechanism has been suggested to explain that. (B) Treatment reduced significantly the percentage of PIL, pneumothorax and apnea as well. But after the treatment some parameters are still high in the WOSS-6 group, this result highlights again that (B) is not absolutely efficient against critical illness. There was a significant variation between (A) & (B) only in WOSS-4 group indicating that (B) is better than (A) in other words (B) is the best treatment promoting a faster efficient recovery among these 3 in a condition the patient should not suffer from a severe stage of COVID-19. According to another study, the results of tocilizumab treatment are inspiring. The temperature of all the patients returned to normal very quickly. The respiratory function and all other symptoms improved remarkably. Tocilizumab treatment is recommended to reduce the mortality of severe COVID-19. [13] Some of our study results deal with their results including the significant decrease in fever even in severe cases. The novel score will mainly include the following parameters: number of comorbidities, age and inflammation biomarkers since they were the most correlated ones to severity leading to mortality.

## 5 Conclusion

This study reports the association between the primary risk factors which are the number of comorbidities & age, and the increased risk of mortality in COVID-19 patients. Thereafter, we explored the variation in the blood test’s biomarkers including mainly LDH, CRP, D-dimer, IL-6 levels & the lymphocyte and neutrophils count, and variation in the percentage of pulmonary inflammatory lesions which revealed a high inflammation in COVID-19 patients in the severe stages of this disease. All these data suggest that an elder person suffering from more than two comorbidities, with high level of inflammatory biomarkers is at high risk of developing critical CoV disease. Finally, the efficiency of many treatments including antivirals and immunosuppressive medicine was examined and our results indicated that Tocilizumab exhibited a decrease in the inflammation biomarkers and in the percentage of pulmonary inflammatory lesions more than antivirals and anti-retrovirals even if it wasn’t totally efficient against severe cases of COVID-19.

This study provides a look into the underlying processes that lead to mortality in this disease which may offer new targets in the treatment of this pandemic which had caused millions of confirmed deaths, making it one of the deadliest in history. Another treatment could be a combination between Tocilizumab and an anti-inflammatory molecule capable of decreasing other inflammatory markers. Moreover, the most significant biomarkers associated with severe cases of COVID-19 represent potential biomarkers of great importance (components of a novel score) that can provide early prediction of cases needing admission to an intensive care unit, mechanical ventilation, and an early prediction of death. Perhaps this novel score will be adopted in the Lebanese hospitals and perhaps mortality will be reduced after delivering proper treatment according to each case.

## Data Availability

All data produced in the present study are available upon reasonable request to the authors

---

**Supplementary table 1:**
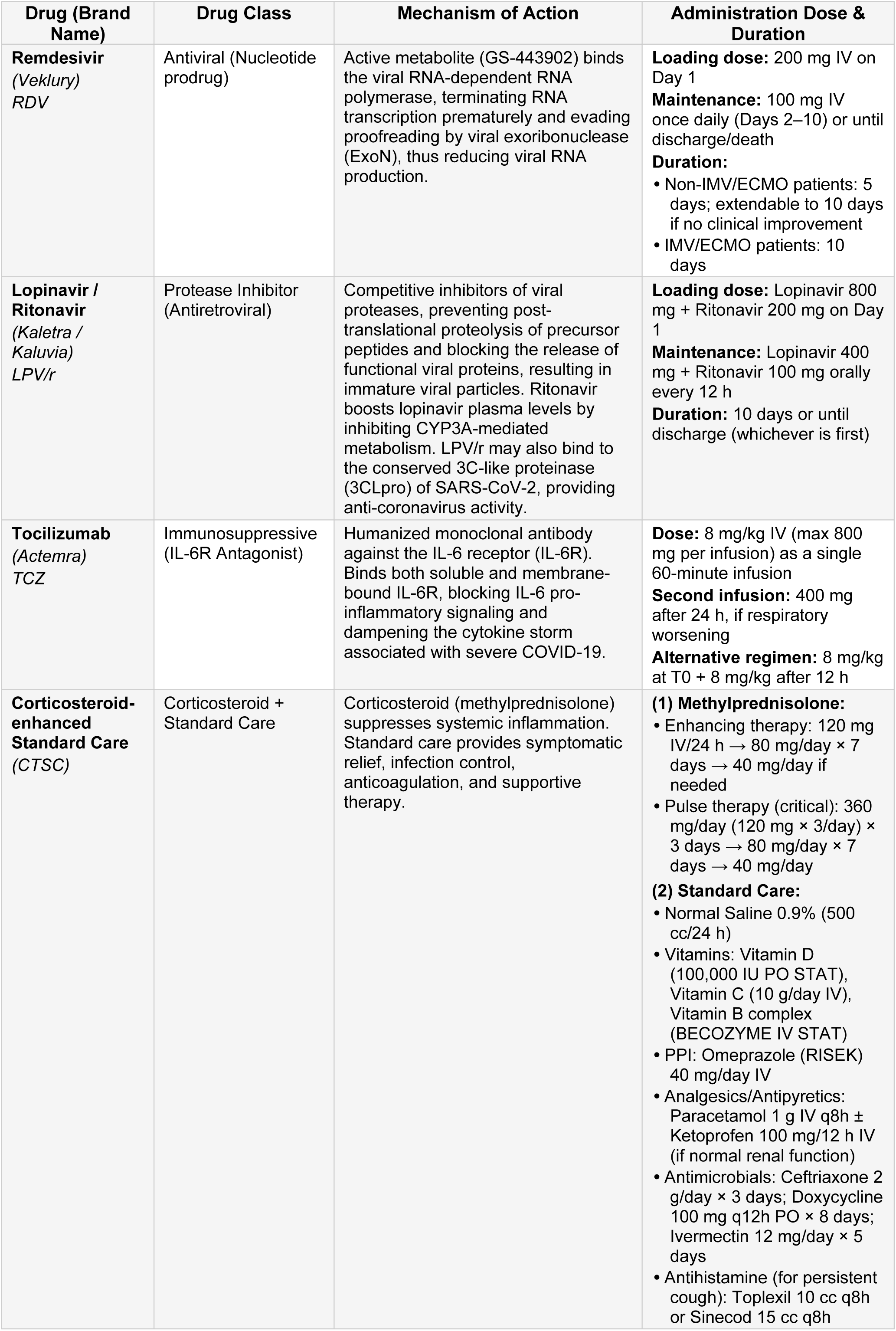
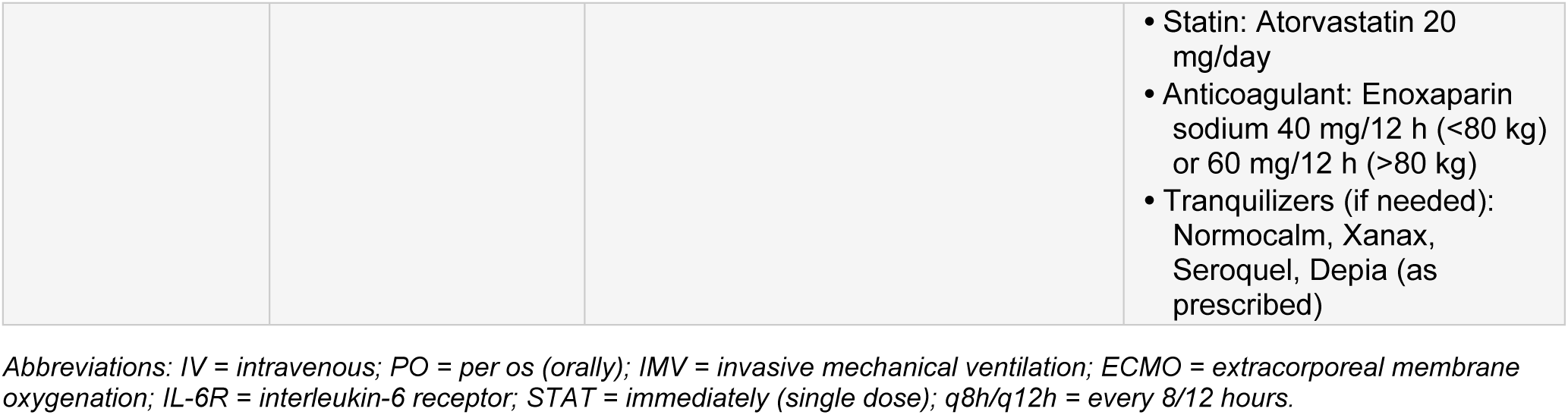
Treatment Protocols — COVID-19 Study.

